# Left Atrial Fibrosis Amelioration Following Cardioversion Assessed with LGE-MRI in Patients with Persistent Atrial Fibrillation

**DOI:** 10.1101/2025.09.29.25336934

**Authors:** Fang Qin, Yunlin Chen, Tianli Xia, Zheng Fang, Danni Liu, Weixiang Song, Wenjiang Chen, Huaan Du, Na Wang, Peilin Xiao, Weijie Chen, Bo Zhang, Junan Chen, Chunxia Gan, Pei Zhou, Xinting Zhu, Haitao Ran, Dajing Guo, Zhiyu Ling, Yuehui Yin

## Abstract

**Background:** Previous studies have demonstrated that cardioversion of atrial fibrillation may alleviate atrial remodeling. We aimed to investigate whether left atrial (LA) tissue fibrosis assessed by LGE-MRI, may be reversed after conversion of persistent atrial fibrillation (PersAF) to sinus rhythm.

**Methods:** Patients with PersAF underwent two LGE-MRI scans following cardioversion were prospectively recruited. LA fibrosis was categorized into core zone (IIR > 1.61) and border zone (0.97<IIR≤1.61) based on the image intensity ratio (IIR). Reverse remodeling of LA was defined as at least a 15% reduction in the max LA volume (LAV). The differences in baseline and follow-up LA structure, function and quantified fibrosis were compared. Multivariable analysis was used to assess the associations of border and core zone changes with other LA parameters.

**Results:** Fifty-three patients were included. The area of the border [22.2 (11.8-37.0) vs. 18.1 (11.8-25.2) cm^2^, *p*= 0.037] and core zones [0.9±3.0 vs. 0.1±0.6 cm^2^, *p*= 0.010] decreased from baseline, along with reductions in LAV, increase of LA ejection fraction and strain. After multivariable adjustment, ΔLATEF was correlated with Δborder zone (r^2^=0.155, *p*= 0.015), baseline left ventricular mass and ΔLATEF was correlated with Δcore zone (r^2^=0.286, *p*<0.001). The cardiac reverse remodeling is not related with Δborder zone and Δcore zone.

**Conclusions:** Atrial fibrosis improved concomitantly with LA shrinkage and enhanced LA function after PersAF to sinus rhythm. LA fibrotic reverse is an independent parameter compared with the traditional definition of cardiac reverse remodeling. The clinical implications warrant further investigation.

## Introduction

Atrial fibrillation (AF) is a prevalent form of atrial tachyarrhythmia and a major contributor to stroke, heart failure, and cognition impairment, placing a substantial burden on both public health and the economy. Notably, AF is both a consequence and a causal factor in the remodeling of the left atrium (LA) ^1^. Over time, AF tends to progress towards a more persistent form ^2^, with fibrosis being a key pathological hallmark of the atrial substrate. This has been supported by histological studies involving surgical specimens, autopsy findings, magnetic resonance imaging (MRI), and electroanatomic voltage mapping ^3,4^. These studies have demonstrated that LA scarring correlates with significant LA enlargement and has been identified as a predictor of AF recurrence ^5^, highlighting the complex pathological mechanisms underlying persistent atrial fibrillation (PersAF) substrates.

Treatment options for PersAF, including pharmacological interventions, direct current cardioversion, and catheter ablation, often yield less favorable outcomes compared to those for paroxysmal AF. Despite significant advancements in ablation techniques and strategies, high rates of recurrence persist among patients with PersAF who underwent catheter ablation ^6^. Current guidelines emphasize the need to assess risk factors for AF recurrence—such as left atrial size, patient age, AF duration, and comorbidities—prior to ablation ^7,8^.

Restoring sinus rhythm may interrupt the vicious cycle of AF initiation and persistence, potentially prevent the progression of cardiovascular remodeling and even facilitate the reversal of certain remodeling features. These include increased atrial refractoriness, reduced LA size, and improved atrial function as assessed by myocardial strain analysis using cardiac echocardiography and cardiac magnetic resonance (CMR) ^9^. However, dynamic evaluation of LA fibrosis changes remains understudied in this context. Traditional pathological methods have proven unsuitable for such evaluations, but MRI with late gadolinium enhancement (LGE-MRI) has emerged as a promising tool. LGE-MRI provides valuable insights into cardiac structure, function, tissue characteristics, and fibrosis quantification in the LA. Additionally, LGE-MRI allows for serial assessments, which may rival traditional pathological examinations^9-11^.

Despite these technological advances, the reversibility of atrial fibrotic tissue following AF termination remains inconclusive. Therefore, the primary aim of this study is to investigate the dynamic reversibility of LA fibrosis by utilizing LGE-MRI in patients undergoing electrical or pharmacological cardioversion for PersAF—procedures that do not directly impact LA fibrosis. This approach may offer new insights into the management of PersAF.

## Materials and Methods

### Study Design

This study focused on patients with PersAF who underwent late gadolinium-enhanced magnetic resonance imaging (LGE-MRI) immediately and 3-6 months after achieving sinus rhythm restoration. The collected information encompassed demographic characteristics, comorbidities, symptoms, laboratory and imaging findings, drug prescriptions, as well as details regarding electrical cardioversion (ECV) procedures. Our study adhered to the principles outlined in the Declaration of Helsinki guidelines. Ethical approval was obtained from the medical ethics committee of the second affiliated hospital of Chongqing medical university, and informed consent was obtained in accordance with the International Ethical Guidelines for Health-Related Research Involving Humans ^10^. This study has been registered in www.clinicaltrials.org, and received identifier of NCT05099783.

### Study Population

Inclusion Criteria Eligible patients were those diagnosed with PersAF and enlarged LA (i.e., a continuous episode lasting more than 7 days, and LA diameter was 38-55 mm which measured by transthoracic echocardiography) who underwent successfully sinus rhythm restoration through either electrical or pharmacological cardioversion, and sinus rhythm was maintained by administrating antiarrhythmic agents. The first CMR evaluation was conducted 1-3 days after cardioversion and the next during the follow-up of 3-6 months. All patients received breathing training before CMR examination.

Exclusion Criteria Patients with the following diseases or conditions were excluded from the study: (1) Valvular heart disease; a history of previous interventions, including radiofrequency ablation for AF, atrial septal defect occlusion, or complex congenital heart disease surgery, hypertrophic cardiomyopathy, primary dilated cardiomyopathy; (2) a history of myocardial infarction, unstable angina pectoris, syncope, or cerebrovascular accidents within the past six months; (3) patients with cardiac pacemaker; (4) uncontrolled hyperthyroidism; (5) poor quality of images due to displacement of the chest, frequent premature and uncontrolled breathing movement during CMR scanning.

### CMR Protocol

CMR images were acquired using 3.0 T scanners (MAGNETOM Prisma, Siemens Healthcare, Erlangen, Germany) equipped with electrocardiographic (ECG) and respiratory gating and a 16-channel body phased array coil. Subjects were positioned in a supine posture. Cardiac two-, three-, four-chamber views, and short-axis cine images encompassing the entire left ventricle (LV) and LA were obtained using the balanced steady-state free precession sequence (b-SSFP) with the following imaging parameters: repetition time (TR) = 37.56ms, echo time (TE) = 1.38ms, field of view (FOV) = 400×335mm, matrix size = 208×139, flip angle = 51°, temporal resolution = 46 to 60 ms, slice thickness = 8 mm, and slice gap = 1.6mm.

Late gadolinium enhancement (LGE) images were captured 15 to 20 minutes following an intravenous bolus of gadolinium-DOTA (Gadoterate Meglumine) at a dose of 0.2 mmol/kg. This was accomplished using a three-dimensional (3D) inversion recovery gradient-echo pulse sequence with respiratory navigation and ECG gating (atrial end-diastole, determined from four-chamber cine). Typical acquisition parameters were as follows: voxel size = 1.5×1.5×2.5 mm, TR = 651ms, TE = 1.22 ms, flip angle = 15°, bandwidth = 814 Hz/pixel, and TI = 270-320 ms. A TI scout sequence (Look-Locker sequence) within a mid-ventricular image was utilized to nullify the normal myocardial signal and establish the optimal inversion time for LGE-MRI. The mean acquisition time for a 3D-LGE sequence was 15 minutes (ranging from 11 to 20 minutes), depending on heart rate and breathing patterns. The slice thickness = 2.5mm.

All patients underwent imaging examinations at baseline (T0) and 3-6 months postoperatively (T1). The ΔLA border zone was defined as the difference between the area at T1 and that at T0 (Δ = T1 - T0), where a negative value indicates a reduction in area.

### LA fibrosis Analysis

LA LGE-MRI data was transferred to a workstation (ADAS, Galgo Medical SL, Barcelona, Spain) for image analysis. The image intensity ratio (IIR), a previously described LGE-RMI technique, was calculated. IIR is defined as the mean pixel intensity of each sector divided by the mean pixel intensity of the entire LA blood pool. It was categorized as follows: IIR < 0.97 for normal myocardium, 0.97<IIR≤1.61 for border zone fibrosis, and IIR > 1.61 for dense fibrosis/scar^11^,^12^ ensuring interpatient comparability of LA fibrosis. LA regions were segmented and manually defined by tracing epi- and endocardial contours in each axial plane by an investigator blinded to the clinical outcomes. To reduce endocardial and epicardial artifacts as well as partial volume effects, a 3-dimensional (3D) shell representing the mid-atrial wall (50% thickness) was automatically constructed by ADAS and manually adjusted based on MRI images. Pulmonary veins and the mitral valve were excluded from fibrosis analysis. Pixel signal intensity (SI) maps, automatically calculated from LA DE-MRI, were projected into the atrial mid-myocardial shell, color-coded, and presented in histograms. The volume of segmented LGE enhancement was determined by summing the segmented areas of enhancement in each axial slice. The LA LGE volume, as quantitated above, was divided by the total LA tissue volume to calculate an LA LGE percentage. Figure 1 illustrates the segmented LA by tracing epi- and endocardial contours in each axial plane, with color-coded zones representing 0.97<IIR≤1.61 and IIR > 1.61 in the 3D model.

**Figure 1.**
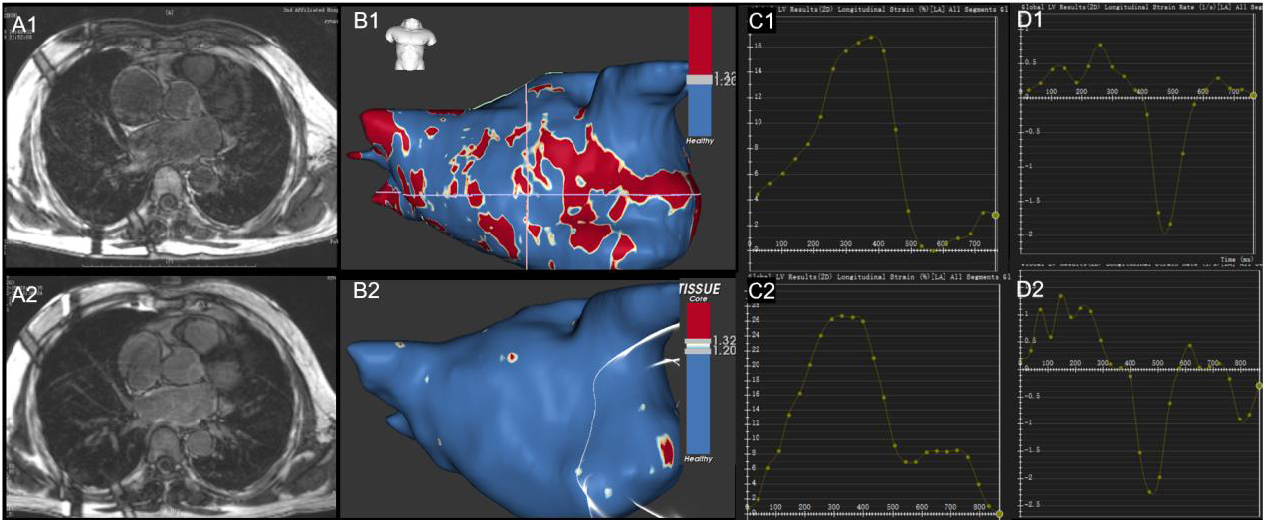
The segmented LA with color-coded zones representing border and core zone in the 3D model and LA strain and strain rate assess. A1: the segmented LA by tracing contours in each axial plane, B1: color-coded zones representing border zone (0.97 < IIR ≤ 1.61, yellow) and core zone (IIR > 1.61, red) and normal zone (IIR ≤ 0.97, blue) in the 3D model. A2: the reexamination of segmented LA for the patient, B2: the reexamination of color-coded zones in the 3D model for the patient. C1: the LA strain assessed by MRI; C2: the reexamination of LA strain for the patient; D1: the LA strain rate assessed by MRI; D2: the reexamination of LA strain rate for the patient.

### LA Volumetric and Functional Analysis

LA volumetric and functional parameters were analyzed using CVI42 software (version 5.0, Circle Cardiovascular Imaging Inc., Calgary, Canada) by two radiologists with 5- and 10-year experience in CMR imaging, respectively. Left atrial cyclic volumes, including LAV_max_ (defined as LAV at LV end-systole before mitral valve opening), LAV min (defined as LAV at LV end-diastole and before mitral valve closing), and LAV p-ac (defined as LAV at LV mid-diastole before LA contraction), were derived from individual time-volume curves using Simpson’s method on short-axis cine images. All volumetric parameters were indexed to body surface area.

From the LAV measurements, LA total/passive/active emptying fractions (EF) were calculated using the following formulas: (1) LA total EF (LATEF) = (LAV_max_ – LAV_min_) / LAV_max_ × 100%, (2) LA passive EF (LAPEF) = (LAV_max_ – LAV_p-ac_) / LAV_max_ × 100%, (3) LA active EF (LAAEF) = (LAV_pre-a_ – LAV_min_) / LAV_p-ac_ × 100%. LA length parameters were measured at LV end-systole, using the frame immediately before the opening of the mitral leaflets in the 2-, 3-, and 4-chamber views. The sphericity index was calculated as the ratio of LA maximum volume to the volume of a sphere with the maximum LA length diameter among atrial length and transverse length (perpendicular to the atrial length) from the 2- and 4-chamber images. Reverse remodeling of LA was defined as at least a 15% reduction LAV_max_^13^.

Left atrial sphericity (LASP) was calculated. The segmented LA model consists of triangles defined by 3D segmentation points. The center of mass was determined first for the model. Then, the average radius (AR) from this center of mass was calculated as the average of radii of all model triangles, weighted by triangle areas. The best-fitted sphere was defined as a sphere with its center at the center of mass and a radius of AR. The coefficient of variation of the AR weighted by triangle area was calculated (coefficient of variation of the sphere, CVS), and from this, LASP was calculated as (1 - CVS) * 100%. This method uses the square root of summed squared differences in calculation, resulting in lower values compared to our 3DS method^14^.

### LA Myocardial Strain Analysis

CMR-Feature tracking (CMR-FT) analysis was conducted on the acquired b-SSFP cine images using CVI42 software (Version 5.0, Circle Cardiovascular Imaging Inc., Calgary, Canada) by two independent radiological experts blinded to other MRI information and clinical data. The fundamentals of LA strain algorithms have been previously described, and their validity has been established^15^. In summary, a set of cine images in the three long-axis views (2-, 3-, and 4-chamber) was imported into the feature tracking module. The endocardial contours of LA myocardium were manually delineated using a point-and-click approach at end-diastole and end-systole, with subsequent automatic tracking throughout the cardiac cycle (excluding pulmonary veins and LA appendage). The quality of automatic tracking was verified and manually adjusted if necessary. Longitudinal strain was defined as the temporal variation of the LA contour length and was calculated as (L(t) - L0) / L0, where L0 is the initial length, and L(t) is the length at a given time. The radial motion fraction was defined as the radial relative displacement toward the LA center of mass and was calculated as (M(t) - M0) / M0, where M0 is the initial radius, and M(t) is the radius at a given time. Three aspects of LA strain were analyzed as previously described: total strain (εs, corresponding to LA reservoir function), active strain (εa, corresponding to LA booster pump function), and passive strain (εe, corresponding to LA conduit function, defined as the difference between εs and εa). Accordingly, three strain rate (SR) parameters were evaluated: peak positive SR (SRs, corresponding to reservoir function), peak early negative SR (SRe, corresponding to conduit function), and peak late negative SR (SRa, corresponding to booster pump function). Tracking was repeated three times, and the LA strain and SR values were derived from the average of the three repeated measurements. Figure 1 illustrates the analysis process of global LA all-segment longitudinal strain and strain rate.

### Statistical Analysis

The normality of quantitative variables was assessed using the one-sample Kolmogorov-Smirnov test. Continuous variables are presented as mean ± standard deviation (SD) for normally distributed data and as median (P25-P75) for non-normally distributed data. Categorical variables are expressed as frequencies with percentages: n (%). For comparisons of variable levels, one-way ANOVA was used for normally distributed data, while the Mann-Whitney U test was applied for non-normally distributed data. The chi-square test was used to analyze the distribution of categorical variables. A two-sided P-value < 0.05 was considered statistically significant. For comparing differences between paired continuous variables during follow-up, the paired t-test was used when the differences between the two groups approximately followed a normal distribution and the Wilcoxon signed-rank test was used as an alternative when this normality assumption was not met. Multivariable linear regression models were constructed to evaluate the association between demographic factors and changes in CMR parameters during the follow-up periods after cardioversion. Logistic regression analysis was used to identify associations between CMR structure and function variables and LA reverse remodeling. All statistical analyses were performed using SPSS 19.0 software (SPSS Inc., Chicago, IL, USA) for Windows.

## Results

Atrial tissue fibrosis estimation via LGE-MRI was successfully quantified twice in 60 PersAF patients after they were converted to sinus rhythm. Seven patients (11.7%) were excluded due to poor CMR quality.

### Baseline Clinical Characteristics

Fifty-three patients were included. The mean age was 64.3 years old and 81.1% was male, with AF durations with 12.0 (1.0-48.0) months. The average CHA2DS2-VASc score was 2.0 (1.0-3.0).

The demographic characteristics and comorbidity profiles did not differ between the reversed group and the non-reversed group at baseline.

According to the change of LA volume, seventeen of them who exhibited a decrease in LAV of more than 15% were included in reversed group and 36 patients in the non-reverse remodeling group after 3-month follow-up. The baseline clinical characteristics were similar between the two groups. The average age for patients with reversed LA seemed younger than the non-reversed but there was no significant importance (61.1±9.5 vs. 65.9±8.6 yrs, *P*=0.076). The AF duration in reversed LA group seemed shorter than the non-reversed but there was no statistical significance[6.0 (1.0-42.0) vs. 12.0 (1.8-48.0) months, *p*=0.449]. The baseline clinical data were displayed in table 1.

**Table 1.**
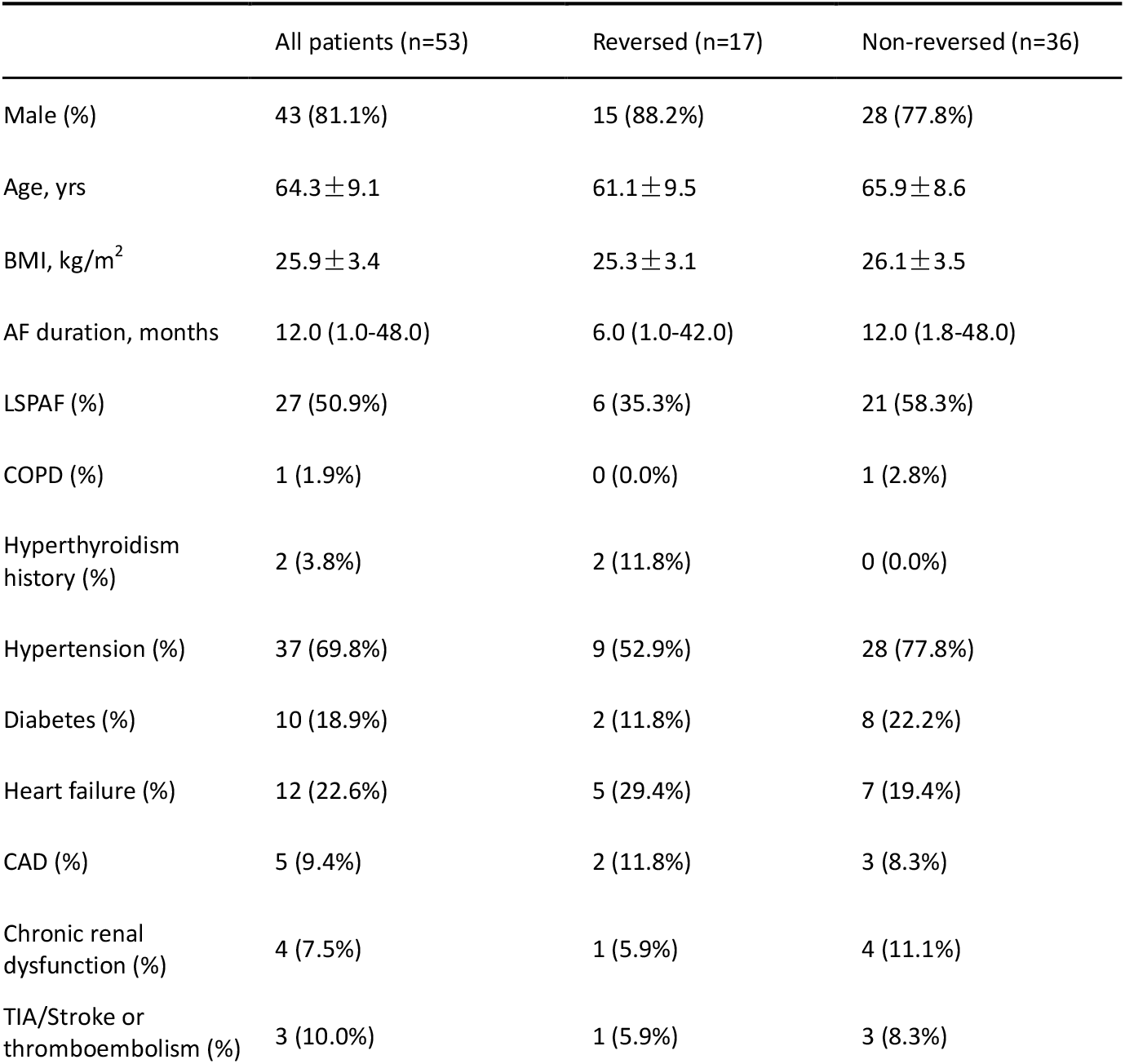
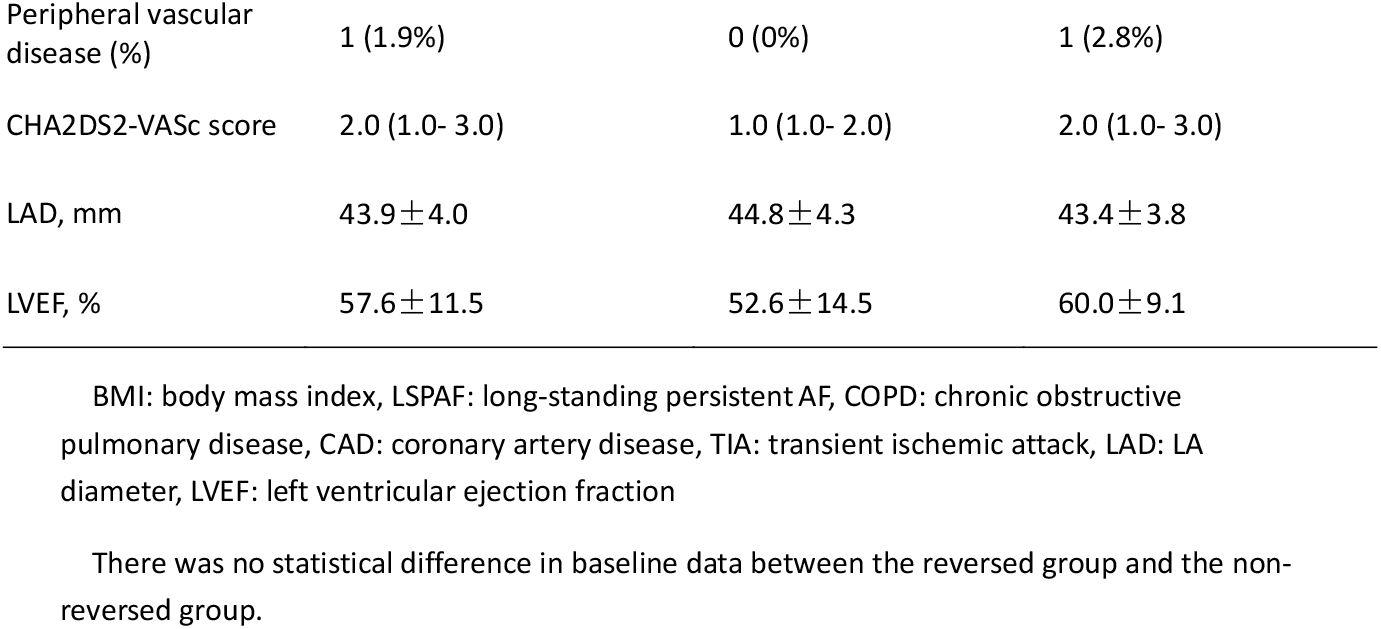
Clinical Characteristics of Study Cohort at baseline.

|  | All patients (n=53) | Reversed (n=17) | Non-reversed (n=36) |
| --- | --- | --- | --- |
| Male (%) | 43 (81.1%) | 15 (88.2%) | 28 (77.8%) |
| Age, yrs | 64.3 ± 9.1 | 61.1 ± 9.5 | 65.9 ± 8.6 |
| BMI, kg/m<sup>2</sup> | 25.9 ± 3.4 | 25.3 ± 3.1 | 26.1 ± 3.5 |
| AF duration, months | 12.0 (1.0-48.0) | 6.0 (1.0-42.0) | 12.0 (1.8-48.0) |
| LSPAF (%) | 27 (50.9%) | 6 (35.3%) | 21 (58.3%) |
| COPD (%) | 1 (1.9%) | 0 (0.0%) | 1 (2.8%) |
| Hyperthyroidism history (%) | 2 (3.8%) | 2 (11.8%) | 0 (0.0%) |
| Hypertension (%) | 37 (69.8%) | 9 (52.9%) | 28 (77.8%) |
| Diabetes (%) | 10 (18.9%) | 2 (11.8%) | 8 (22.2%) |
| Heart failure (%) | 12 (22.6%) | 5 (29.4%) | 7 (19.4%) |
| CAD (%) | 5 (9.4%) | 2 (11.8%) | 3 (8.3%) |
| Chronic renal dysfunction (%) | 4 (7.5%) | 1 (5.9%) | 4 (11.1%) |
| TIA/Stroke or thromboembolism (%) | 3 (10.0%) | 1 (5.9%) | 3 (8.3%) |
| Peripheral vascular disease (%) | 1 (1.9%) | 0 (0%) | 1 (2.8%) |
| CHA2DS2-VASc score | 2.0 (1.0- 3.0) | 1.0 (1.0- 2.0) | 2.0 (1.0- 3.0) |
| LAD, mm | 43.9 ± 4.0 | 44.8 ± 4.3 | 43.4 ± 3.8 |
| LVEF, % | 57.6 ± 11.5 | 52.6 ± 14.5 | 60.0 ± 9.1 |

BMI: body mass index, LSPAF: long-standing persistent AF, COPD: chronic obstructive pulmonary disease, CAD: coronary artery disease, TIA: transient ischemic attack, LAD: LA diameter, LVEF: left ventricular ejection fraction
There was no statistical difference in baseline data between the reversed group and the non-reversed group.

### CMR Characteristics

Compared to the baseline, the average border zone area was decreased[22.2 (11.8-37.0) vs. 18.1 (11.8-25.2) cm^2^, *p*=0.037], and the same trend for core zone (0.9±3.0 vs. 0.1±0.6cm^2^, *p*=0.010). The core zone percentage decreased (0.7±2.3 vs. 0.1±0.5%, *p*=0.010) after cardioversion. The border zone percentage decreased but there was no significant difference (17.6 (10.5-30.0) vs. 16.9 (10.0-24.6) %, *p*=0.351).

Several LA variables significantly improved after cardioversion, including LAVI (60.3±19.6 vs. 53.6±16.6 ml/cm^2^, *p*=0.003), LAV_max_ (*p*=0.001), LAV_min_ (*p*<0.001) and LAV_pre-a_ (*p*<0.001). LA functions were improved in LATEF (32.2±13.1 vs. 44.1±15.8%, *p*<0.001), LAAEF (*p*<0.001) and LAPEF (*p*=0.042). The total strain (18.2±8.6 vs. 27.4±14.1%, *p*<0.001), and active strain and passive strain were improved totally (*p*<0.001 for both). The LA peak positive strain rate (0.8±0.4 vs. 1.2±0.6s^-1^, *p*<0.001) and LA peak late negative strain rate (−0.7±0.6 vs. −1.3±0.9s^-1^, *p*<0.001) were improved. No significant differences were observed in LA peak early negative strain rate. Left ventricular ejection fraction improved after several months under sinus rhythm (57.5±11.5 vs. 61.4±8.4%, *p*=0.012). Figure 1 shows the reduction in atrial fibrotic area as visualized via three-dimensional reconstruction. For details, refer to Table 2 and Figure 2.

**Table 2.** CMR Characteristics of Study Cohort at baseline and follow-up.

|  | Cardioversion treatment (n=53) |  | *p* value |
| --- | --- | --- | --- |
|  | Baseline | Follow-up |  |
| LVEF, % | 57.5 ± 11.5 | 61.4 ± 8.4 | 0.012* |
| Border zone, cm<sup>2</sup> | 22.2 (11.8- 37.0) | 18.1 (11.8- 25.2) | 0.037* |
| Core zone, cm<sup>2</sup> | 0.9 ± 3.0 | 0.1 ± 0.6 | 0.010* |
| Border zone ratio, % | 17.6 (10.5- 30.0) | 16.9 (10.0- 24.6) | 0.351 |
| Core zone ratio, % | 0.7 ± 2.3 | 0.1 ± 0.5 | 0.010* |
| LASP | 81.0 ± 3.9 | 80.8 ± 4.0 | 0.518 |
| LAV<sub>max</sub>, ml | 109.4 ± 32.7 | 97.2 ± 26.3 | 0.001* |
| LAV<sub>min</sub>, ml | 76.7 ± 32.5 | 57.3 ± 28.3 | <0.001* |
| LAV<sub>pre-a</sub>, ml | 81.2 (63.9- 98.6) | 99.8 (65.8- 108.3) | <0.001* |
| LAVI, ml/m<sup>2</sup> | 60.3 ± 19.6 | 53.6 ± 16.6 | 0.003* |
| LATEF, % | 32.2 ± 13.1 | 44.1 ± 15.8 | <0.001* |
| LAPEF, % | 17.1 ± 7.7 | 19.0 ± 9.3 | 0.042* |
| LAAEF, % | 18.1(7.6- 25.0) | 31.3 (18.9- 39.9) | <0.001* |
| LA εs, % | 18.2 ± 8.6 | 27.4 ± 14.1 | <0.001* |
| LA εe, % | 11.9 ± 6.3 | 16.4 ± 7.5 | <0.001* |
| LA εa, % | 6.3 ± 4.4 | 11.0 ± 8.0 | <0.001* |
| LA SRs, s<sup>-1</sup> | 0.8 ± 0.4 | 1.2 ± 0.6 | <0.001* |
| LA SRe, s<sup>-1</sup> | -1.2 ± 0.7 | -1.3 ± 0.7 | 0.089 |
| LA SRa, s<sup>-1</sup> | -0.7 ± 0.6 | -1.3 ± 0.9 | <0.001* |

LVEF: left ventricle ejection fraction, LASP: left atrial sphericity, LAV: left atrium volume, LAV<sub>max</sub>: LAV at LV end-systole before mitral valve opening, LAV<sub>min</sub>: LAV at LV end-diastole and before mitral valve closing, LAV<sub>pre-a</sub>: LAV at LV mid-diastole before LA contraction, LAVI: left atrial volume index, LATEF: LA total EF, LAAEF: LA active EF; LAPEF: LA passive EF; εs: total strain, εe: passive strain, εa: active strain, SRs: peak positive strain rate, SRe: peak early negative strain rate, SRa: peak late negative strain rate.
\*: $p < 0.05$

**Figure 2.**
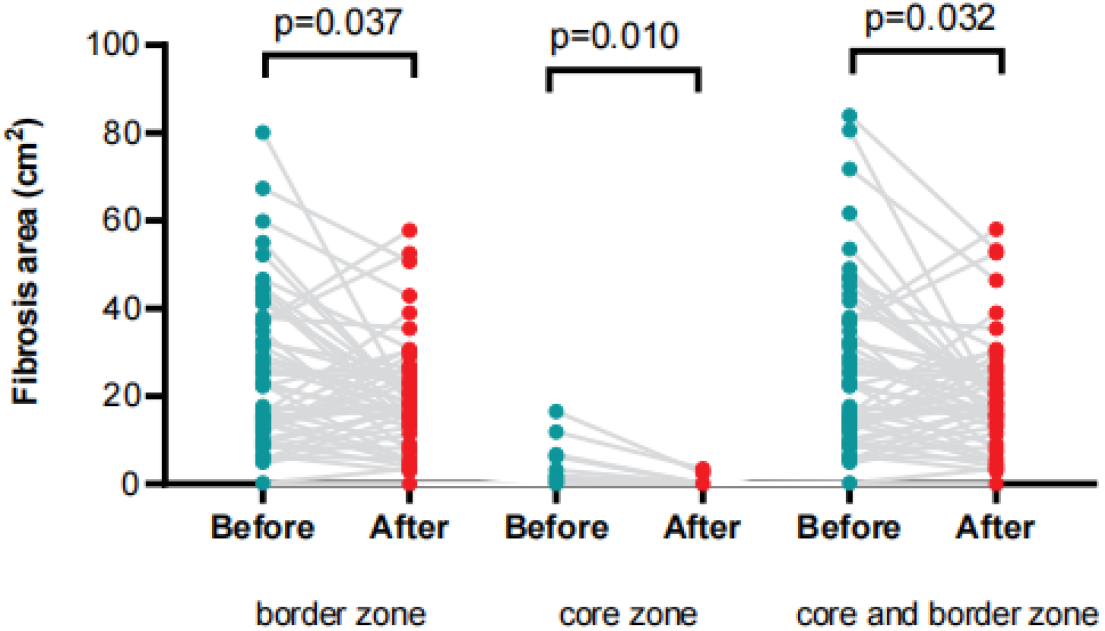
Changes in left atrial fibrotic area at baseline and follow-up. **What is Known** •AF drives LA remodeling, progresses to PersAF, and sinus rhythm may reverse LA structure and function remodeling, but atrial fibrosis reversibility post-AF termination is unclear. **What the Study Adds** •LA fibrosis improved concomitantly with LA shrinkage and enhanced LA function. •LA fibrotic reverse is an independent parameter relative to the traditional definition of cardiac reverse remodeling that defined by a ≥15% reduction in LA volume.

### CMR Characteristics in Reversed and Non-reversed Group

At baseline, in the reversed group, there was a larger LAV_max_[120.8 (96.4-152.7) vs. 110.4 (80.8-123.6), *p*<0.001], LAVI[64.4 (58.0-82.4) vs. 57.0 (39.7-68.5), *p*=0.050], LA border zone [36.7 (15.9-48.2) vs. 16.0 (11.7-31.2), *p*=0.037]and core zone[0 (0-2.5) vs. 0 (0-0), *p*=0.012] when compared with the non-reversed group. The LASP[78.5 (77.0-80.6) vs. 83.3 (78.7-84.7), *p*=0.026]in reversed group was lower. The extent of LATEF (18.5±16.0 vs. 8.6±10.7%, *p*=0.008) and LAAEF (18.8±19.3 vs. 10.7±11.8%, *p*=0.049) improvement were higher in reversed group. However, there was no differences between reversed and non-reversed group in ΔLAPEF (*p*=0.087), ΔLA εs (*p*=0.364), ΔLA εe (*p*=0.443) and ΔLA εa (*p*=0.508). Furthermore, the ΔLA border zone (−12.5±18.2 vs. 61.4±8.4%, *p*=0.027) and Δcore zone (*p*=0.033) were different between reversed and non-reversed group, meaning that the reduction in the area of the LA border zone and core zone were greater in reversed group. There was no difference between reversed and non-reversed group in ΔLASP. Refer to Table 3 for details.

**Table 3.** CMR Characteristics of Reversed and Non-reversed group.

|  | Reversed (n=17) | Non-reversed (n=36) | *p* value |
| --- | --- | --- | --- |
| LAV reduction percentage, % | 28.8 (19.1-38.4) | 2.9 (3.3-6.9) | < 0.001* |
| LAV<sub>max</sub>, ml | 120.8 (96.4-152.7) | 110.4 (80.8-123.6) | 0.045* |
| ΔLAV<sub>max</sub>, ml | -28.5 (-47.5- -21.8) | -3.7 (-7.8- 3.0) | < 0.001* |
| LAVI, ml/m<sup>2</sup> | 64.4 (58.0-82.4) | 57.0 (39.7-68.5) | 0.05 |
| ΔLAVI, ml/m<sup>2</sup> | -21.3 ± 10.1 | 0.4 ± 9.1 | < 0.001* |
| LATEF, % | 27.7 ± 13.2 | 34.4 ± 12.6 | 0.085 |
| ΔLATEF, % | 18.5 ± 16.0 | 8.6 ± 10.7 | 0.008* |
| LAAEF, % | 14.9 ± 12.7 | 19.9 ± 11.7 | 0.065 |
| ΔLAAEF, % | 18.8 ± 19.3 | 10.7 ± 11.8 | 0.049* |
| LAPEF, % | 15.3 ± 6.0 | 18.4 ± 7.9 | 0.266 |
| ΔLAPEF, % | 4.2 ± 7.5 | 0.3 ± 5.3 | 0.087 |
| LA εs, % | 15.4 (10.7-20.8) | 16.7 (13.4-23.4) | 0.181 |
| ΔLA εs, % | 11.6 ± 14.2 | 8.1 ± 12.1 | 0.364 |
| LA $\epsilon$ e, % | 11.3 (5.9-13.5) | 11.8 (7.2-14.9) | 0.407 |
| ΔLA εe, % | 5.7 ± 8.1 | 3.9 ± 7.5 | 0.443 |
| LA $\epsilon_a$, % | 4.7 (2.7-8.0) | 6.4 (3.0-9.2) | 0.344 |
| ΔLA $\epsilon_a$, % | $5.9 \pm 9.4$ | $4.2 \pm 6.6$ | 0.508 |
| Border zone, cm<sup>2</sup> | 36.7 (15.9-43.5) | 15.8 (11.8-30.6) | 0.041* |
| ΔBorder zone, cm<sup>2</sup> | $-12.5 \pm 18.2$ | $-2.0 \pm 14.3$ | 0.027* |
| Border zone reduction percentage, % | $10.2 \pm 14.2$ | $2.0 \pm 13.0$ | 0.036* |
| Core zone, cm<sup>2</sup> | 0 (0-2.5) | 0 (0-0) | 0.012* |
| ΔCore zone, cm<sup>2</sup> | 0 (-2.5-0) | 0 (0-0) | 0.033* |
| Core zone reduction percentage, % | $3.1 \pm 3.0$ | $0.2 \pm 0.8$ | 0.031* |
| Border and core zone, cm<sup>2</sup> | 36.7 (15.9-48.2) | 15.8 (11.8-30.6) | 0.034* |
| ΔBorder and core zone, cm<sup>2</sup> | $-14.5 \pm 19.6$ | $-2.2 \pm 14.9$ | 0.015* |
| Border and core zone reduction percentage, % | $11.7 \pm 15.2$ | $2.1 \pm 13.4$ | 0.020* |
| LASP | 78.5 (77.0-80.6) | 83.3 (78.7-84.7) | 0.026* |
| ΔLASP | 0.5 (-1.6-1.4) | 0.5 (-1.0-1.5) | 0.568 |

LAV: left atrium volume, LAV<sub>max</sub>: LAV at LV end-systole before mitral valve opening, LAVI: left atrial volume index, LATEF: LA total EF, LAAEF: LA active EF; LAPEF: LA passive EF; $\epsilon_s$ : total strain, $\epsilon_e$ : passive strain, $\epsilon_a$ : active strain, LASP: Left atrial sphericity, $\Delta Y = Y_{\text{post}} - Y_{\text{pre}}$

### Factors Associated with Changes in Fibrosis and Left Atrial Remodeling Status

Multiple linear regression was employed to predict ΔBorder zone based on LASP and ΔLATEF, and to predict ΔCore zone based on left ventricular mass (LVmass) and ΔLAV_max_. The regression model for ΔBorder zone was statistically significant (*p*=0.015, adjusted R^2^=0.122). The variable ΔLATEF included in the model had a statistically significant effect on ΔBorder zone (β=-0.275, *p*=0.041). The regression model for ΔCore zone was also statistically significant P<0.001, adjusted R^2^=0.258). Both independent variables included in the model, namely LVmass (β=-0.309, *p*=0.013) and ΔLAV_max_ (β=0.401, *p*=0.002), had statistically significant effects on ΔCore zone.

Binary logistic regression was used in this study to evaluate the impact of LAV_max_, LASP, ΔLATEF, ΔBorder zone, and ΔCore zone on whether the subjects achieved LA reverse remodeling. The resulting logistic model is statistically significant, with χ^2^=25.85 and *p*<0.001. Among the 5 independent variables included in the model, LAV_max_, LASP at the baseline and ΔLATEF is statistically significant. For every 1-unit improvement in LATEF, the possibility of cardiac reverse remodeling increases by 14.1%; for every 1-unit increase in baseline LA volume, the possibility of cardiac reverse remodeling increases by 3.6%; for every 1-unit increase in baseline LASP, the possibility of cardiac reverse remodeling decreases by 34%. The possibility of cardiac reverse remodeling is not related to the improvement degree of cardiac fibrosis. Detailed results are presented in Table 4.

**Table 4.**
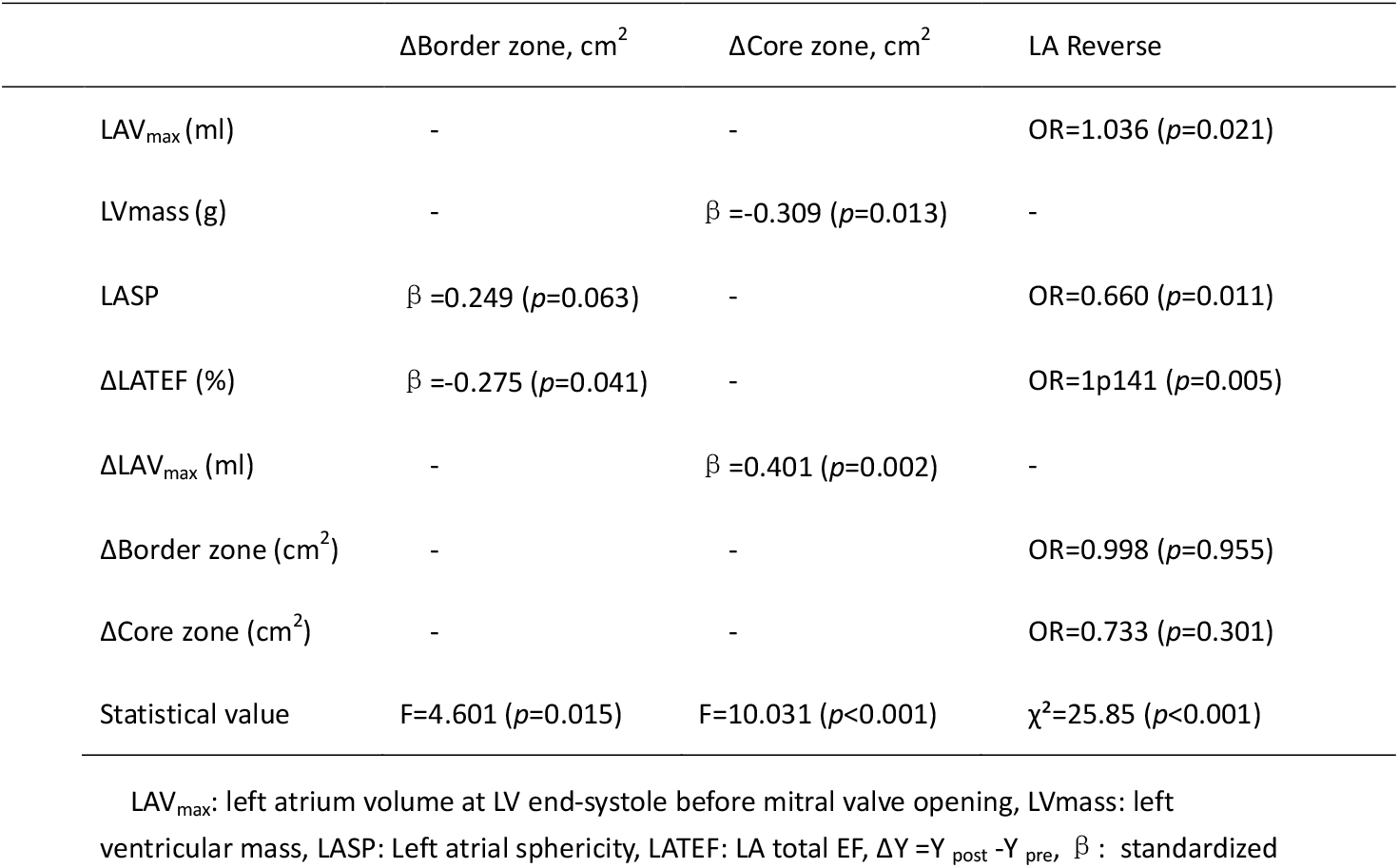
Factors Associated with Changes in Fibrosis and Left Atrial Remodeling Status ΔBorder zone, cm^2^ ΔCore zone, cm^2^ LA Reverse.

## Discussion

AF induces extensive remodeling of the LA, involving functional, electrical, metabolic, neurohormonal, and structural changes. Cardioversion interrupts the tachycardic process and may reverse these pathophysiological alterations. This study is the first to dynamically assess LA fibrous tissue in human subjects using LGE-MRI, demonstrating the reversal of LA fibrosis following sinus rhythm restoration in patients with PersAF. This highlights the adaptability of atrial tissue, which can respond positively to therapeutic interventions, indicating the potential for reversing pathological changes.

Beyond the reversal of LA fibrotic remodeling, our study also reveals improvements in a broader spectrum of parameters, including LA volume, LAEF, and LA strain, underscoring the critical role of maintaining sinus rhythm in preserving atrial health. The data show that a greater increase in LATEF correlates with a larger reduction in the border zone area. Moreover, a larger baseline LVmass and a more substantial reduction in LAV_max_ are associated with a more significant reduction in the core zone. Increased LATEF, together with a large baseline LAV_max_ and a small LASP, were associated with a higher likelihood of LA reverse remodeling-defined as a ≥15% reduction in LAV_max_. However, atrial fibrosis changes were not correlated with this remodeling process.

Atrial LGE-MRI employs IIR to differentiate between distinct tissue zones within the atrial wall. As previously reported, the core zone represents dense, non-viable tissue, indicative of complete fibrosis, while the border zone is a transitional area with intermediate signal intensity, reflecting heterogeneous tissue with partial fibrosis and viable myocytes. This zone harbors slow-conduction channels, which are potential substrates for arrhythmias^16^. Remodeling of the border zone correlates with functional improvements in the LA, whereas changes in the core zone are more closely linked to structural alterations in both the LV and LA. This suggests distinct mechanisms underlying functional versus structural reverse remodeling of the LA. Those atrial subvital myocytes exhibit the potential for reversibility, accompanied by reduced atrial myocardial fibrosis and improved atrial function—similar to hibernating myocardium following myocardial infarction, which can undergo reversible functional restoration after revascularization^17,18^.

Currently, commonly used markers of LA reverse remodeling primarily focus on changes in LAV, neglecting other functional and structural improvements in the atria. LGE-MRI is gold standard for LAV and LA function assessment^19^ and can be used as a surrogate for assessing atrial fibrosis, has proven reliable for providing crucial insights into the location and extent of fibrotic tissue in a non-invasive and repeatable manner^20^. MRI techniques extend beyond diagnostic applications, offering valuable tools for risk stratification and prognostic evaluation^21^. This study employs LGE-MRI to precisely investigate not only changes in LA structure and function but also early atrial myocardial fibrosis, thereby facilitating a deeper understanding of subclinical atrial lesions ^22^. All patients in this study underwent MRI under sinus rhythm and trained before MRI scan to ensure optimal image quality. The rate of poor MRI image quality in this study was 11.7%, which was lower than the 17% reported in the DECAAF study, some of which were thought to be resulted from arrhythmia, fast heart rate and irregular respiration^21^.

Restoration of sinus rhythm effectively rescues reversible myocytes to avoid sliding into an irreversible abyss^23^. This intervention reduces the risk of cardiovascular events, whether achieved via electrical cardioversion, pharmacological agents^24^, or radiofrequency ablation^25^. Atrial fibrosis plays a pivotal role in initiating and perpetuating AF, and the current consensus suggests that extracellular matrix remodeling, accompanied by apoptosis and fibrosis, is typically irreversible^26^. In contrast, Prabhu et al. demonstrated the regression of diffuse ventricular fibrosis through T1 mapping following sinus rhythm restoration via catheter ablation in AF patients with systolic dysfunction^27^. In the DECAAF study, atrial fibrosis was assessed using CMR before and after radiofrequency ablation for AF. The baseline fibrosis was 18.7 ± 8.7%, with residual fibrosis at 15.8 ± 8.0%. However, the procedures performed in that study [pulmonary vein isolation (PVI) and complex fractionated atrial electrogram (CAFÉ) ablation] induced fibrosis themselves, complicating the accurate assessment of intrinsic fibrosis^28^. In contrast, our study used either pharmacological or electrical cardioversion, which did not affect the atrial substrate, thereby avoiding these confounding factors.

Beyond the reversal of LA fibrotic remodeling, our findings also highlight a broader spectrum of reverse remodeling, including improvements in LA diameter and strain. This aligns with previous studies^29^. Notably, Akoum et al. have demonstrated a correlation between atrial fibrosis and conditions such as atrial appendage thrombus and spontaneous contrast in AF patients, suggesting that severe atrial fibrosis may increase the risk of thrombosis [27, 28]. The benefits of early rhythm control therapy in reducing concomitant cardiovascular events have been well-documented^24,30^. However, further work is required to explore the relationship between fibrotic tissue reduction, functional improvements, and long-term prognostic benefits.

Atrial fibrosis results from a multifactorial interplay of neurohormonal, inflammatory, metabolic, genetic, and environmental factors^31^. The extent and stages of fibrosis correlate with both LAVI and the duration of AF^32^. The degree of atrial fibrosis, as assessed by MRI, serves as a crucial predictor for arrhythmia recurrence following catheter ablation^21,33^. However, when comparing MRI-guided fibrosis ablation combined with PVI to PVI alone, no significant difference was observed in atrial arrhythmia recurrence^34^. In this study, we demonstrated that LA fibrotic tissue, as well as structural and functional changes, can occur following cardioversion. For patients with PersAF, relying solely on baseline fibrosis to assess disease severity may have limited clinical value. In those patients with both structural and functional atrial abnormalities, evaluating whether fibrosis or function can be reversed may offer a more comprehensive assessment of disease status. Recurrence after radiofrequency catheter ablation for PersAF is common, and several clinical, anatomical, and procedural factors have been identified as predictors^35^. The most consistent and strongest predictor is an increased left atrial size or volume. LAV, as measured by CMR, is a robust marker of atrial remodeling, AF recurrence risk, and mortality. Its accurate assessment is critical for guiding clinical decision-making, particularly in the context of ablation and risk stratification. CMR-based LAV measurement is favored due to its precision and prognostic value.

The potential role of irreversible atrial fibrosis may contribute to poor rhythm control in PersAF. Therefore, the reversal of atrial fibrosis could potentially offer a better evaluation of prognosis. Fibrosis changes that occur independently of volume alterations may serve as a valuable prognostic factor. However, further research is necessary to fully explore this hypothesis.

### Study Limitations

Several limitations of our study should be acknowledged. First, the relatively small sample size may limit the generalizability of our findings. Second, the retrospective design, including non-sequential patient recruitment, introduces the potential for biases that could affect the measurement of clinical outcomes. Lastly, while magnetic resonance imaging provides invaluable insights, its high cost and time-consuming nature represent significant barriers to widespread clinical application.

## Conclusions

This study provides compelling evidence for the reversibility of LA fibrosis remodeling and sheds light on the intricate relationship between atrial remodeling and the efficacy of cardioversion in managing AF. These findings underscore the need for continued research into personalized treatment approaches and highlight the importance of refining our understanding of atrial fibrosis and its clinical implications.

## Data Availability

The data referred to in the manuscript are not publicly available due to ethical restrictions related to human participant privacy. However, de-identified datasets with all personal information removed will be made available to qualified researchers upon reasonable request. Requests can be sent to the corresponding author with a brief description of the intended research use.

## Non-standard Abbreviations and Acronyms

AF: atrial fibrillation
PersAF: persistent atrial fibrillation ECV = electrical cardioversion
LA: left atrium
LAV: LA volume
LGE-MRI: magnetic resonance imaging with late gadolinium enhancement
IIR: image intensity ratio

## Sources of Funding

This study is funded partly by Remarkable Innovation – Clinical Research Project, The Second Affiliated Hospital of Chongqing Medical University (No. 2022ZYLCYJ002, Prof. Yuehui Yin). This study was also supported by the National Natural Science Foundation of China (Grant Numbers: 32071110 and Yuehui Yin) and Science-Health Joint Medical Scientific Research Project of Chongqing (Grant Numbers: 2025DBXM007 and Zhiyu Ling).

## Disclosures

All authors report no conflicts of interest.

## Notes

### Competing Interest Statement

The authors have declared no competing interest.

### Clinical Trial

This study has been registered in www.clinicaltrials.org, and received identifier of NCT05099783.

### Author Declarations

The research described in this manuscript was approved by the Ethics Committee of the Second Affiliated Hospital of Chongqing Medical University with the number: 202374

